# Statistical analysis plan for the “Does longer peripheral intravenous catheter Length optimise Antimicrobial Delivery: a randomised controlled trial to reduce interruptions to intravenous antimicrobial delivery (The LEADER Study)”

**DOI:** 10.1101/2023.10.05.23296519

**Authors:** Asmaa El-Heneidy, Robert S. Ware, Catherine O’Brien, Emily Larsen, Nicole Marsh, Amanda Corley

## Abstract

The “Does longer peripheral intravenous catheter (PIVC) Length optimise Antimicrobial Delivery: a randomised controlled trial to reduce interruptions to intravenous antimicrobial delivery (The LEADER Study)”, is a randomised controlled trial (RCT). This RCT aims to assess if long peripheral intravenous catheters (PIVCs) (4.5-6.4 cm) when compared with short-PIVCs (<4 cm) optimise antimicrobial delivery by reducing PIVC failure rates.

The purpose for this document is to minimise bias and ensure transparency and internal validity for the findings of the trial, by defining and making publicly available the analysis approach prior to reviewing or analysing trial data. The statistical analysis plan (SAP) will inform analysis and reporting of the main effectiveness findings of the trial. It provides a detailed description of the primary and secondary trial outcomes and the methods for statistical comparison.

## 1. Administrative information

### 1.2 Study identifiers

- ***Protocol:*** Corley, A., O’Brien, C., Larsen, E., Peach, H., Rickard, C., Hewer, B., Pearse, I., Fenn, M., Cocksedge, R., & Marsh, N. (2023). Does longer peripheral intravenous catheter length optimise antimicrobial delivery? Protocol for the LEADER study. British journal of nursing, 32(7); S24-S30.

https://doi.org/10.12968/bjon.2023.32.7.S24.

- *ANZCTR registration number:* ACTRN12621001199808

## 2. Study synopsis

The LEADER study is a single-centre, two-arm, parallel group RCT, comparing short- and long-PIVCs for patients requiring intravenous (IV) antimicrobial therapy for three days or more.

### 2.1 Study Objectives

1. To compare device failure in long-PIVCs, with usual care (short-PIVCs).
2. To reduce the incidence of interrupted IV antimicrobial administration.
3. To assess the safety profile of long-PIVCs.

### 2.2 Study Hypotheses

#### 2.2.1 PRIMARY HYPOTHESIS

1. Compared to patients receiving usual care (short-PIVC), patients with a long-PIVC will have fewer IV antimicrobial treatment interruptions due to all-cause device failure.

#### 2.2.2 SECONDARY HYPOTHESES

Compared to patients receiving usual care (short-PIVC), those with a long-PIVC will:

1. have fewer vascular access devices inserted over the course of antimicrobial therapy,
2. have significantly fewer central venous access devices (CVAD) inserted,
3. have lower rate of PIVC-related complications (insertion-related complications, bloodstream infection, local infection, phlebitis, occlusion, infiltration/extravasation, thrombosis, and dislodgement).

### 2.3 Eligibility criteria

#### 2.3.1 INCLUSION CRITERIA

- Patients aged 18 years and older.
- Patients who require peripheral IV antimicrobial treatment for a duration of three days or more.
- Patients who can provide informed written consent.

#### 2.3.2 EXCLUSION CRITERIA

- Patients with upper arm limitations (e.g., mastectomy, renal patients).
- Non-English-speaking patients without interpreter.
- Patient receiving end-of-life care.
- Previous enrolment in the study.

### 2.4 Outcomes

#### 2.4.1 PRIMARY OUTCOME PER PROTOCOL: All-cause PIVC failure will be defined as the unplanned removal of the initial catheter during IV antimicrobial therapy: A composite measure of

1. Infiltration/extravasation: The unintended leakage of the IV antimicrobial solution into the interstitial tissue instead of entering the vein properly,
2. Blockage/occlusion: The partial or complete obstruction of the PIVC, resulting in the inability to administer the IV antimicrobial solution. This obstruction can occur with or without external leakage from the site.
3. Phlebitis (defined as pain ≥2/10 on a numerical rating scale; or ≥2 signs/symptoms of pain/tenderness, erythema, swelling, or palpable cord),
4. Suspected or confirmed thrombosis: Suspected or confirmed blood clot formation in or around the PIVC, which can impede the flow of the IV antimicrobial solution (suspected: as assessed/ suspected by the treating clinician; confirmed: ultrasound/ venographic confirmed thrombosed vessel at PIVC site),
5. Dislodgement (complete or partial removal of the PIVC from the vein),
6. Infection: Laboratory-confirmed local or bloodstream infection associated with the PIVC insertion site. This can include both localised infections at the insertion site and systemic infections originating from the PIVC.

#### 2.4.2 SECONDARY OUTCOMES PER PROTOCOL

1. Incidence of All-cause PIVC failure (initial device) per 1000/catheter days.
2. Individual PIVC complications as listed above in the primary clinical outcome.
3. Dwell time of each initial IV study device inserted.
4. Number of first insertion success of initial (randomised) device (yes/no).
5. Number of PIVCs unable to be successfully inserted.
6. Number of insertion attempts (needle punctures) for initial (randomised) device.
7. Adverse events (e.g., nerve damage, adverse skin event [rash, blister, itchiness, skin tears, adhesive residue]).
8. Number and incidence of IV devices inserted per patient to complete the course of antimicrobials (randomised and any subsequent short-PIVC, long-PIVC, midline, or CVAD).
9. Patient-reported insertion pain (0-10 numerical rating scale).
10. Patient-reported satisfaction of PIVC insertion and removal (0-10 numerical rating scale).
11. Staff overall satisfaction with PIVC device (0-10 numerical rating scale).
12. Cost analysis (in a subset of seven participants per study arm), including costs of treating PIVC-related complications and PIVC reinsertion.

### 2.5 Setting

This RCT was conducted in the general medical and surgical wards of a large quaternary referral hospital, the Royal Brisbane and Women’s Hospital, (>900 beds).

### 2.6 Sample size

The primary outcome of interest in the study was the number and percentage, of the initial devices that experienced a disruption to antimicrobial administration due to all-cause post-insertion PIVC failure. The calculations of the sample size were based on relevant local and international data. These data indicated that approximately 43% of short-PIVCs will have an interruption due to antimicrobials, compared to 24% for long-PIVCs. Consequently, the study aimed to recruit a total of 192 participants, with 91 patients to be assigned to each arm of the study. Additionally, an incremental inclusion of 5 participants per arm was planned to account for potential attrition.

This sample size was carefully designed to equip the study with 80% statistical power. meaning it has a high chance of detecting significant differences between the groups at a significance level of P<0.05.

### 2.7 Study interventions

Details on recruitment are outlined in the participant flowchart (Figure 1). Participants were randomised to receive either:

1. Standard care (control): a short-PIVC less than 4cm length e.g., Insyte Autoguard BC inserted into arm veins, or
2. Intervention: a long-PIVC, 4.5 to 6.4 cm length, e.g., Introcan Safety Deep Access or Insyte Autoguard BC inserted in the upper arm or forearm veins.

### 2.8 Randomisation and blinding

To randomly assign patients to either the short-PIVC or long-PIVC group, a centralised web-based randomisation service was used. This service ensured that the allocation of patients was concealed prior to the randomisation process. The randomisation was conducted in a 1:1 ratio, and the block sizes used for randomisation varied randomly between 4 and 6.

Each participant was given a unique study identification number (ID). Since the nature of the intervention made it impossible to blind patients and clinicians to their assigned group, they were aware of their treatment. However, the statistician analysing the data was unaware of the treatment assignments to ensure unbiased analysis. Additionally, the microbiologist and laboratory staff responsible for determining infection outcomes were blinded to the treatment allocation when assessing the infection results.

### 2.9 PIVC care

All PIVCs were inserted, maintained, and removed following the hospital’s established policy. The insertion procedure was conducted by a skilled Research Nurse (ReN) experienced in PIVC insertion. The clinical staff was responsible for the ongoing maintenance of the PIVCs, and the decision to remove a PIVC was made by the treating medical team.

### 2.10 Data collection

Data was collected from the patients’ medical records and entered the Research Electronic Data Capture (REDCap) database. Each patient was assigned a unique study ID, which was used to link their data to the study. Enrolled participants were visited daily by a research nurse, except on weekends. During these visits, the nurse assessed the site where the PIVC was inserted and collect data for both primary and secondary outcomes. The specific details of these outcomes can be found in Table 1. On weekends when the research nurse was not available to visit the patients, data was collected retrospectively from the patients’ medical records.

**Table 1.**
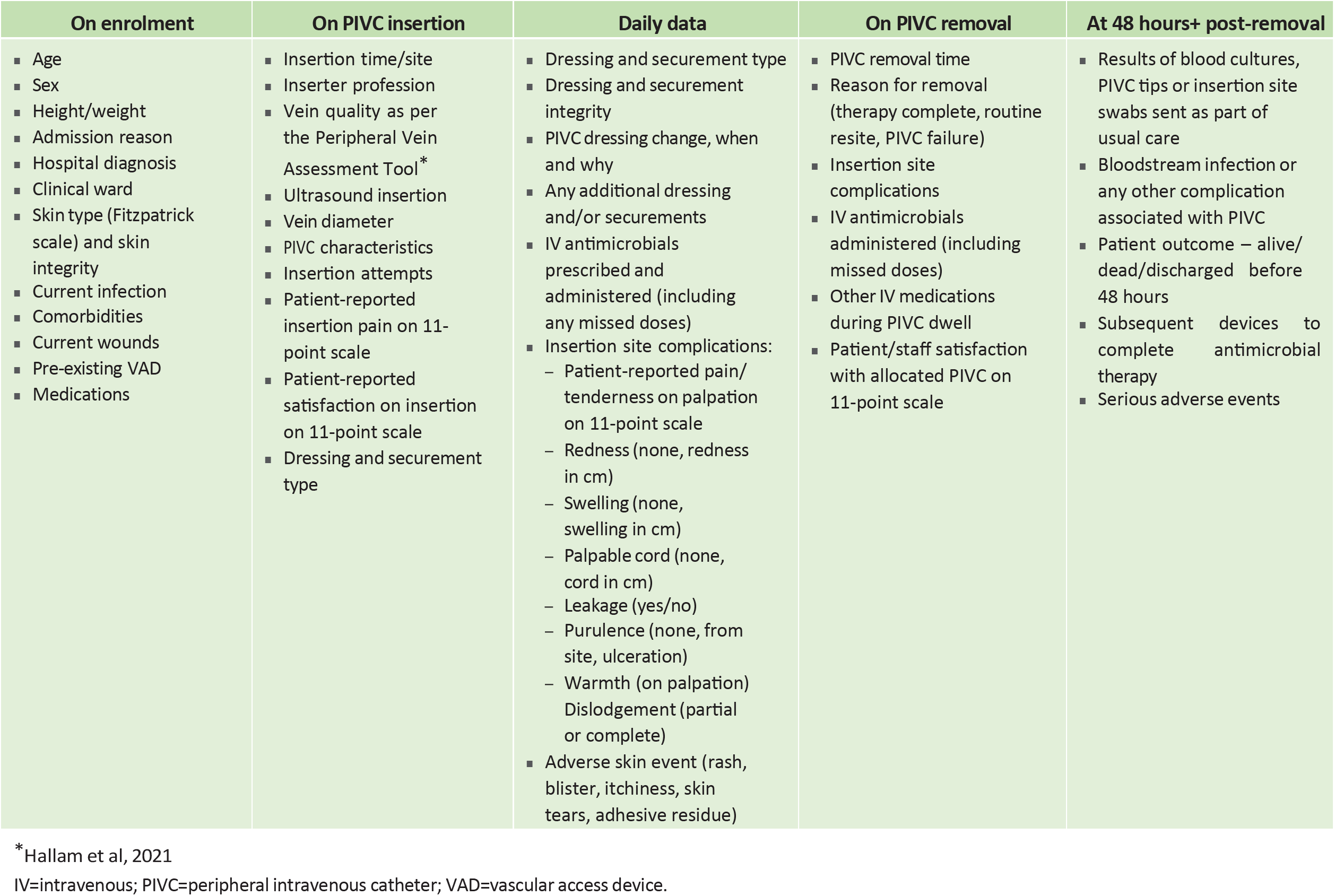
Participant characteristics at baseline.

## 3. Statistical analysis

### 3.1 General principles

#### 3.1.1 INTENTION TO TREAT AND PER-PROTOCOL ANALYSIS

All participants who were randomised and have evaluable data for the endpoint under investigation will be analysed in the study arm to which they were randomised.

For the primary and important secondary outcomes, a per-protocol analysis will be performed to test the sensitivity of the results to noncompliance. For the per-protocol analysis, only participants that received their originally allocated treatment, and who provide outcome data according to protocol, will be included.

The number of missing observations for each primary and secondary outcome will be reported by study group. If the percentage of missing observations is non-trivial (>5%), consideration will be given to the possible introduction of compliance and attrition bias. The cause of any missing data will be assessed. A sensitivity analysis will be conducted to explore the potential impact of missing data. Multiple imputation techniques will be employed as part of this analysis if deemed appropriate.

#### 3.1.2 DATA CLEANING APPROACH

Data collection and quality management will be completed through use of the REDCap database.

#### 3.1.3 PRESENTATION OF RESULTS

Continuous data will be summarised in a descriptive manner using either the mean and standard deviation (SD) or the median and interquartile range (IQR), depending on the distribution characteristics of the variable being examined. Categorical data will be presented as frequencies and percentages.

#### 3.1.4 LEVEL OF SIGNIFICANCE

A significance level of alpha = 0.05 (two-tailed) will be used to evaluate statistical significance.

#### 3.1.5 STATISTICAL SOFTWARE

All estimates will be derived using Stata v14.1 (StataCorp, College Station, Texas, USA).

### 3.2 Evaluation of demographic and baseline characteristics

#### 3.2.1 PARTICIPANT DEMOGRAPHIC AND CLINICAL CHARACTERISTICS

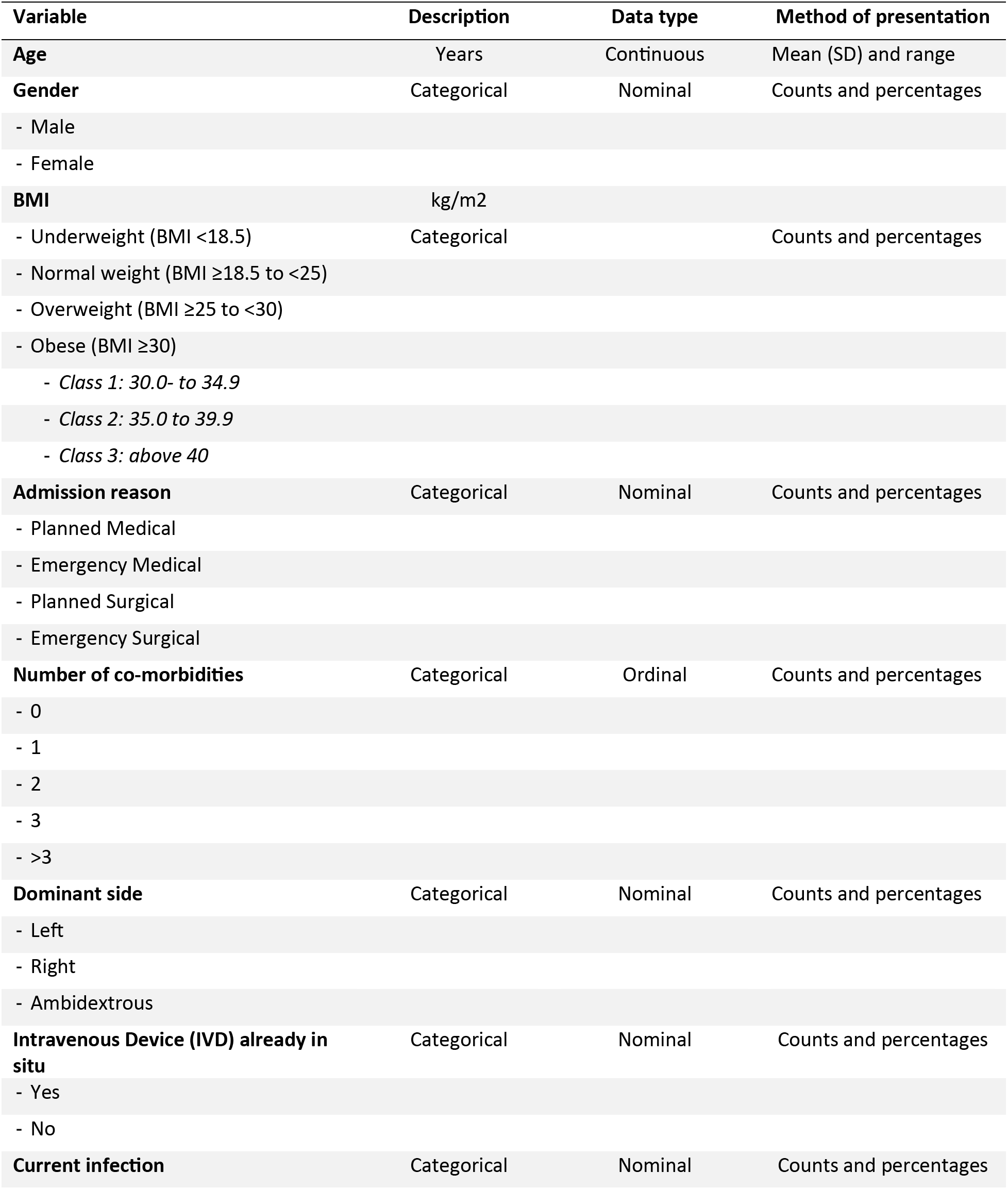
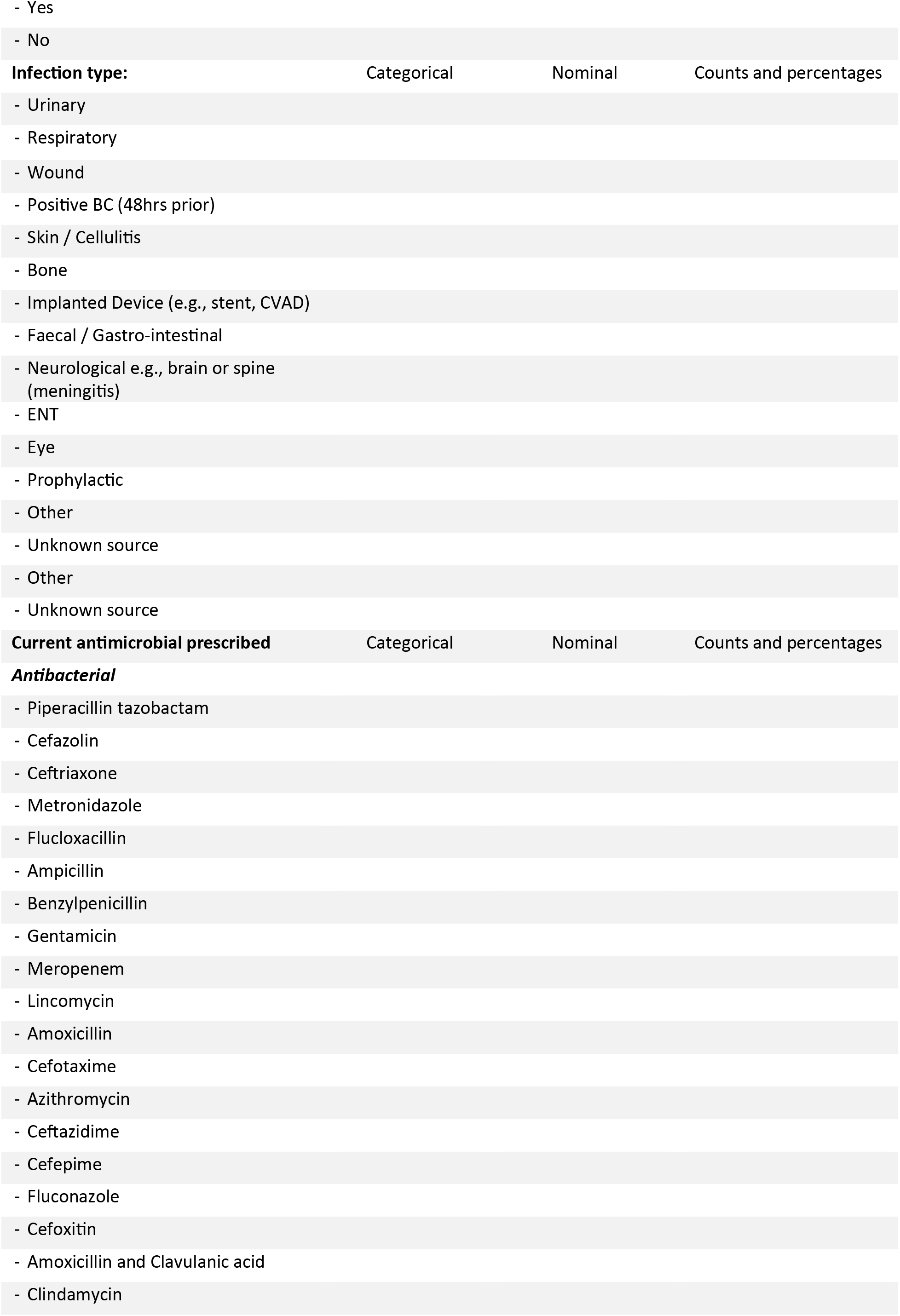
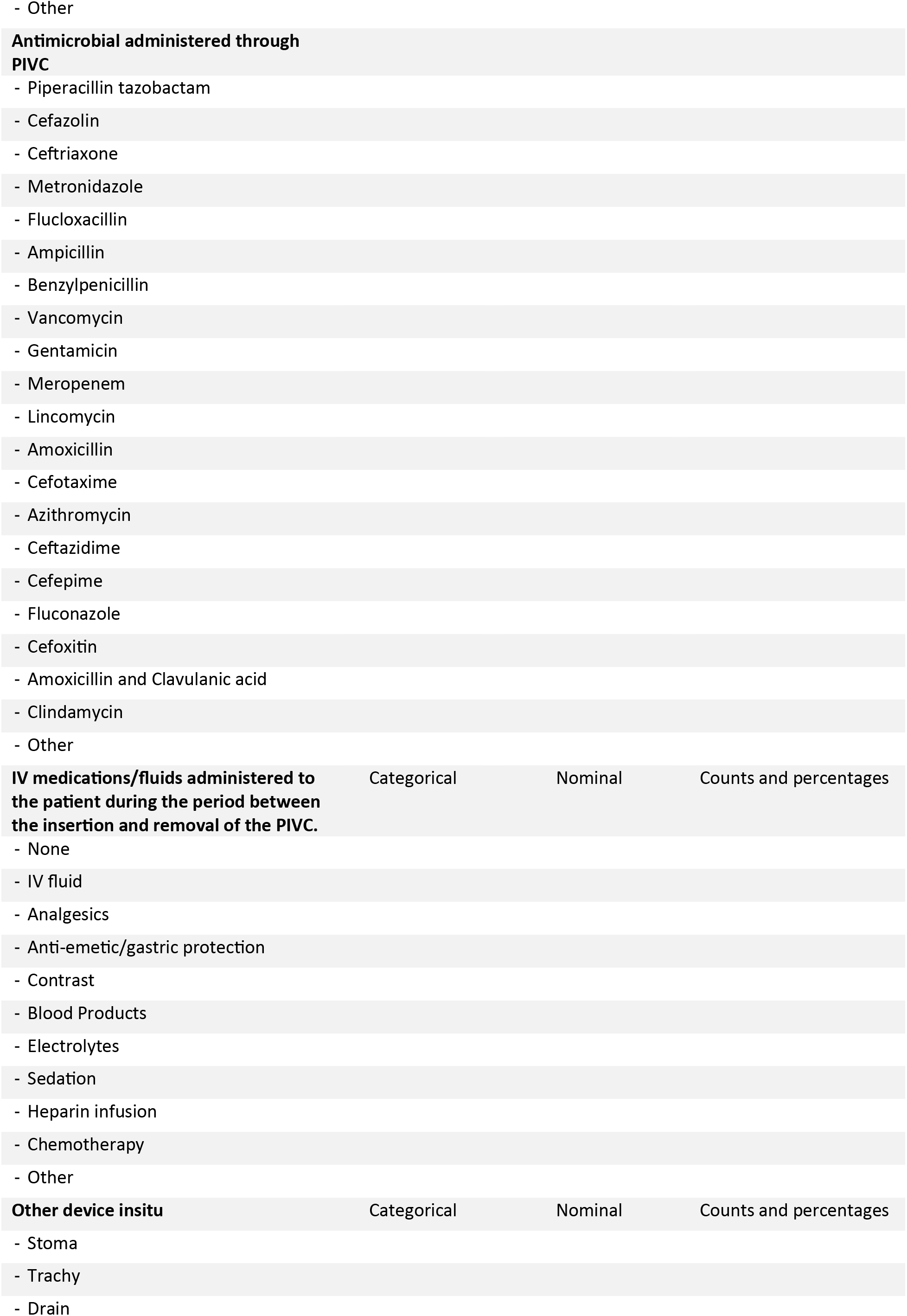
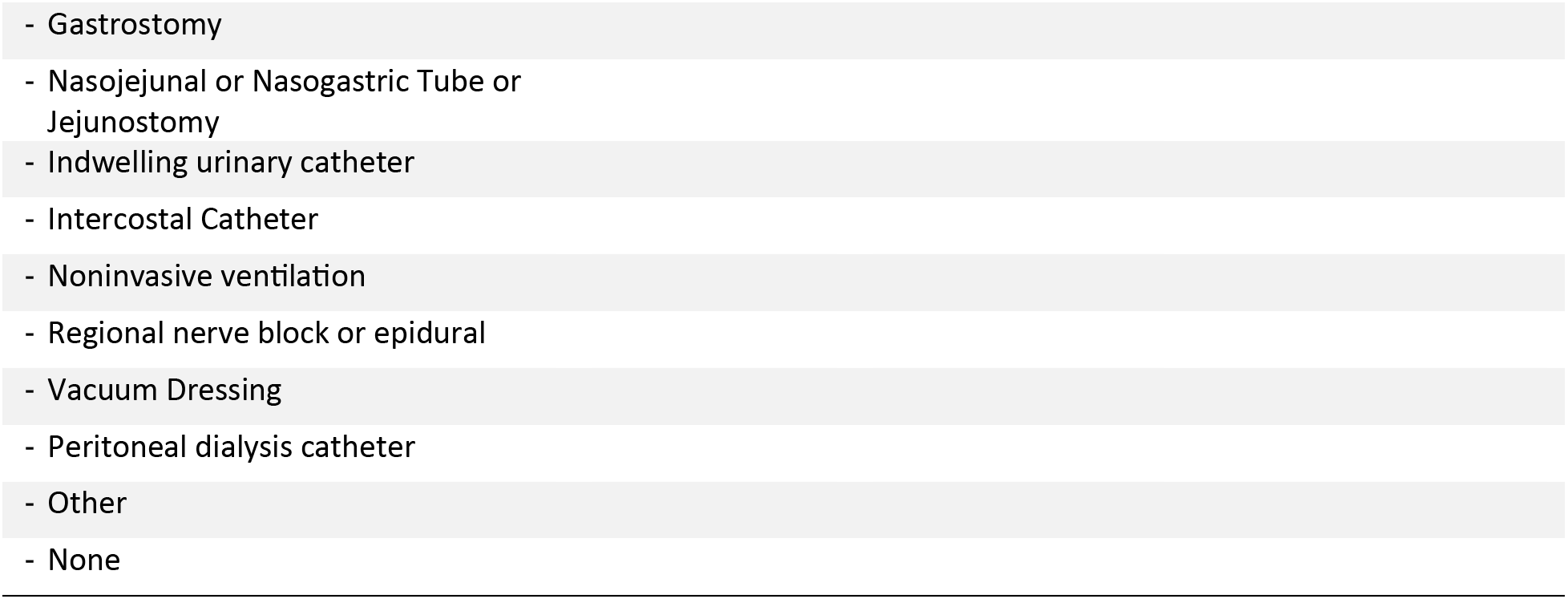
Demographic and clinical data will be reported by allocated study group.

#### 3.2.2 DEVICE CHARACTERISTICS

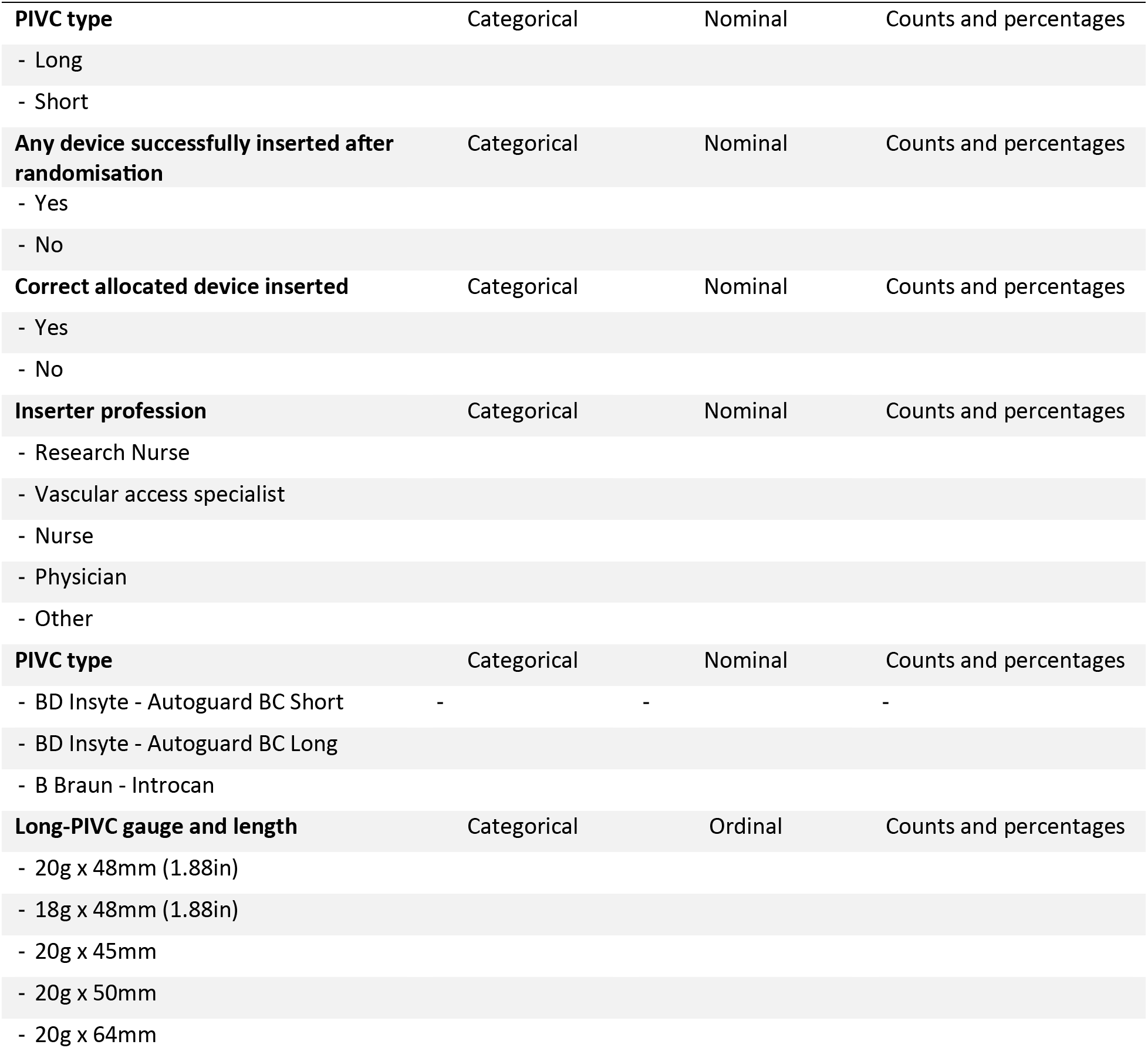
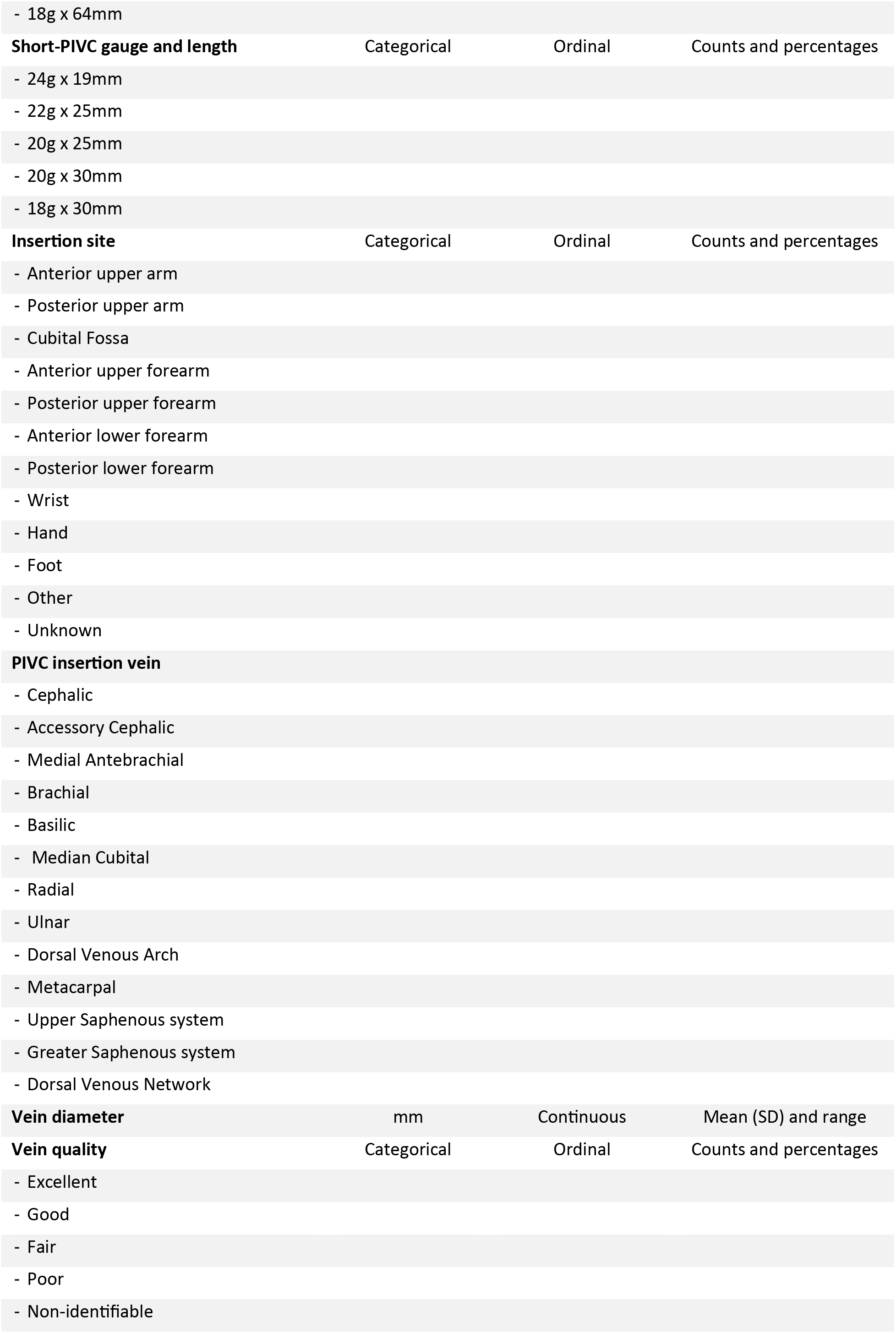
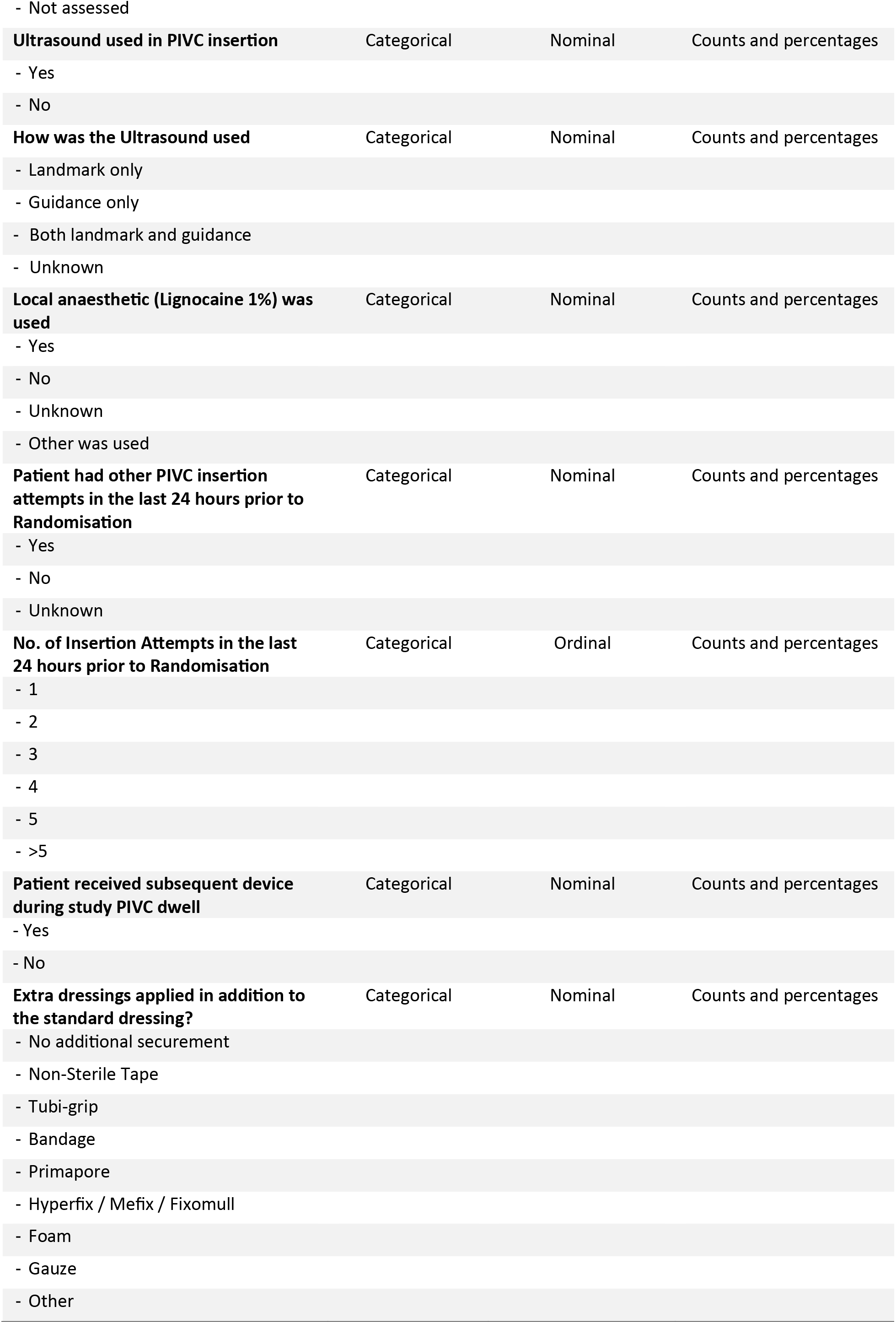
Device characterisCcs collected at baseline will be reported by allocated study group.

**Device characteristics collected at baseline will be reported by allocated study group.**
|  |  |  |  |
| --- | --- | --- | --- |
| **PIVC type** | Categorical | Nominal | Counts and percentages |
| - Long |  |  |  |
| - Short |  |  |  |
| **Any device successfully inserted after randomisation** | Categorical | Nominal | Counts and percentages |
| - Yes |  |  |  |
| - No |  |  |  |
| **Correct allocated device inserted** | Categorical | Nominal | Counts and percentages |
| - Yes |  |  |  |
| - No |  |  |  |
| **Inserter profession** | Categorical | Nominal | Counts and percentages |
| - Research Nurse |  |  |  |
| - Vascular access specialist |  |  |  |
| - Nurse |  |  |  |
| - Physician |  |  |  |
| - Other |  |  |  |
| **PIVC type** | Categorical | Nominal | Counts and percentages |
| - BD Insyte - Autoguard BC Short | - | - | - |
| - BD Insyte - Autoguard BC Long |  |  |  |
| - B Braun - Introcan |  |  |  |
| **Long-PIVC gauge and length** | Categorical | Ordinal | Counts and percentages |
| - 20g x 48mm (1.88in) |  |  |  |
| - 18g x 48mm (1.88in) |  |  |  |
| - 20g x 45mm |  |  |  |
| - 20g x 50mm |  |  |  |
| - 20g x 64mm |  |  |  |
| - 18g x 64mm |  |  |  |
| **Short-PIVC gauge and length** | Categorical | Ordinal | Counts and percentages |
| - 24g x 19mm |  |  |  |
| - 22g x 25mm |  |  |  |
| - 20g x 25mm |  |  |  |
| - 20g x 30mm |  |  |  |
| - 18g x 30mm |  |  |  |
| **Insertion site** | Categorical | Ordinal | Counts and percentages |
| - Anterior upper arm |  |  |  |
| - Posterior upper arm |  |  |  |
| - Cubital Fossa |  |  |  |
| - Anterior upper forearm |  |  |  |
| - Posterior upper forearm |  |  |  |
| - Anterior lower forearm |  |  |  |
| - Posterior lower forearm |  |  |  |
| - Wrist |  |  |  |
| - Hand |  |  |  |
| - Foot |  |  |  |
| - Other |  |  |  |
| - Unknown |  |  |  |
| **PIVC insertion vein** |  |  |  |
| - Cephalic |  |  |  |
| - Accessory Cephalic |  |  |  |
| - Medial Antebrachial |  |  |  |
| - Brachial |  |  |  |
| - Basilic |  |  |  |
| - Median Cubital |  |  |  |
| - Radial |  |  |  |
| - Ulnar |  |  |  |
| - Dorsal Venous Arch |  |  |  |
| - Metacarpal |  |  |  |
| - Upper Saphenous system |  |  |  |
| - Greater Saphenous system |  |  |  |
| - Dorsal Venous Network |  |  |  |
| **Vein diameter** | mm | Continuous | Mean (SD) and range |
| **Vein quality** | Categorical | Ordinal | Counts and percentages |
| - Excellent |  |  |  |
| - Good |  |  |  |
| - Fair |  |  |  |
| - Poor |  |  |  |
| - Non-identifiable |  |  |  |
| - Not assessed |  |  |  |
| **Ultrasound used in PIVC insertion** | Categorical | Nominal | Counts and percentages |
| - Yes |  |  |  |
| - No |  |  |  |
| **How was the Ultrasound used** | Categorical | Nominal | Counts and percentages |
| - Landmark only |  |  |  |
| - Guidance only |  |  |  |
| - Both landmark and guidance |  |  |  |
| - Unknown |  |  |  |
| **Local anaesthetic (Lignocaine 1%) was used** | Categorical | Nominal | Counts and percentages |
| - Yes |  |  |  |
| - No |  |  |  |
| - Unknown |  |  |  |
| - Other was used |  |  |  |
| **Patient had other PIVC insertion attempts in the last 24 hours prior to Randomisation** | Categorical | Nominal | Counts and percentages |
| - Yes |  |  |  |
| - No |  |  |  |
| - Unknown |  |  |  |
| **No. of Insertion Attempts in the last 24 hours prior to Randomisation** | Categorical | Ordinal | Counts and percentages |
| - 1 |  |  |  |
| - 2 |  |  |  |
| - 3 |  |  |  |
| - 4 |  |  |  |
| - 5 |  |  |  |
| - >5 |  |  |  |
| **Patient received subsequent device during study PIVC dwell** | Categorical | Nominal | Counts and percentages |
| - Yes |  |  |  |
| - No |  |  |  |
| **Extra dressings applied in addition to the standard dressing?** | Categorical | Nominal | Counts and percentages |
| - No additional securement |  |  |  |
| - Non-Sterile Tape |  |  |  |
| - Tubi-grip |  |  |  |
| - Bandage |  |  |  |
| - Primapore |  |  |  |
| - Hyperfix / Mefix / Fixomull |  |  |  |
| - Foam |  |  |  |
| - Gauze |  |  |  |
| - Other |  |  |  |

### 3.3 Planned analysis of the primary outcome

The primary outcome of interest is the disruption to antimicrobial administration due to all-cause post-insertion PIVC failure: A composite measure of (infiltration/extravasation, blockage/occlusion, phlebitis, thrombosis, dislodgement, and infection). Summary statistics will be calculated for each study group (patients with short-PIVCs and patients with long-PIVCs). The between-group difference will be examined using a log binomial regression analysis. The outcome will be presented as risk ratio (RR) accompanied by a 95% confidence interval (CI) and a corresponding p-value.

### 3.4 Planned analysis of the secondary outcomes

#### 3.4.1 SECONDARY OUTCOME 1: Incidence of All-cause PIVC failure (initial device) per 1000/catheter days

For each study group, the number and percentage of the initial devices that experience any complication during the dwell time resulting in PIVC failure will be calculated and presented as mean, SD and incidence rate per 1000 catheter days.

Occurrences of PIVC complications will be compared between groups using Poisson regression. The outcome of the Poisson regression will be presented as an incidence rate ratio, accompanied by a 95% CI and a corresponding p-value.

#### 3.4.2 SECONDARY OUTCOME 2: Individual complication types for the initial PIVCs as listed above in the primary clinical outcome (Time to event)

For each study group, the number and percentage of the initial devices that experience any complication during the dwell time will be calculated and presented as mean, SD and incidence rate per 1000 catheter days.

Occurrences of PIVC complications will be treated as time-to-event outcomes. This outcome will be compared between groups as a time-to-event outcome using Cox regression. The Cox regression model will include the study group as a fixed effect. The outcome of the Cox regression will be presented as a hazard ratio, accompanied by a 95% CI and a corresponding p-value.

#### 3.4.3 SECONDARY OUTCOME 3: Dwell time of initial IV study device inserted

For each group, the mean and SD of the dwell time, which represents the duration in hours between the insertion and removal of the initial study device will be calculated. These calculations will be conducted for both groups.

To assess the between-group difference, linear regression will be used with the study group considered as the main effect. This analysis will provide the mean difference, accompanied by a 95% CI and a corresponding p-value.

If the normality assumption of linear regression is not met (i.e., residuals of the linear model not normally distributed), an alternative approach will be implemented. The median and interquartile range (IQR) of the number of IV devices and dwell time will be calculated for each group. Subsequently, a quantile regression analysis will be performed with the study group as the main effect. The results will be presented as the median difference, along with a 95% CI and a p-value.

#### 3.4.4 SECONDARY OUTCOME 4: Overall first insertion success for initial study device (yes/no)

The number and percentage of successful study PIVC device insertion will be calculated for each study group. The between-group difference will be examined using a log binomial regression analysis. The analysis will be conducted on an "intention-to-treat" basis. The outcome will be presented as an RR along with a 95% CI and a corresponding p-value.

#### 3.4.5 SECONDARY OUTCOME 5: Number of insertion attempts for initial study PIVCs (needle punctures)

The mean and SD of the total numbers of skin punctures to attempt PIVC placement for first device will be calculated for both groups. Between group comparison of number of PIVC insertion attempts will use linear regression with study group as main factor. The primary analysis will be ‘intention-to-treat’. The effect estimate presented will be mean difference with a 95% CI and a p-value accompanied by a 95% CI and a corresponding p-value.

If the normality assumption of linear regression is not met, an alternative approach will be implemented. The median and interquartile range (IQR) of the number of IV devices and dwell time will be calculated for each group. Subsequently, a quantile regression analysis will be performed with the study group as the main effect. The results will be presented as the median difference, along with a 95% CI and a p-value.

#### 3.4.6 SECONDARY OUTCOME 6: Adverse events

The number and percentage of patients that experience adverse events caused by PIVCs, including nerve damage, extravasation injury, PIVC related BSI or adverse skin event (rash, blister, itchiness, skin tears, adhesive residue, during the dwell time of the initial PIVCs will be calculated per group. The between-group difference will be examined using a log binomial regression analysis. The outcome will be presented as an RR accompanied by a 95% CI and a corresponding p-value.

#### 3.4.7 SECONDARY OUTCOME 7: Number of IV devices inserted to complete the course of antimicrobials (randomised and any subsequent short-PIVC, long-PIVC, midline catheters or CVAD)

The mean and SD of the total number of IV devices, including the initial device, used by each patient to complete the antimicrobial therapy course will be calculated in each group.

To assess the between-group difference, mixed-effects linear regression will be used with the study group considered as the main effect. This analysis will provide the mean difference, accompanied by a 95% CI and a corresponding p-value.

If the normality assumption of linear regression is not met (i.e., residuals of the linear model not normally distributed), an alternative approach will be implemented. The median and interquartile range (IQR) of the number of IV devices and dwell time will be calculated for each group. Subsequently, a quantile regression analysis will be performed with the study group as the main effect. The results will be presented as the median difference, along with a 95% CI and a p-value.

#### 3.4.8 SECONDARY OUTCOME 8: Incidence of IV devices inserted to complete the course of antimicrobials (per antibiotic days) (randomised and any subsequent short-PIVC, long-PIVC, midline catheters or CVAD)

Incidence of IV devices inserted to complete the course of antimicrobials will be compared between groups using Poisson regression. The outcome of the Poisson regression will be presented as an incidence rate ratio, accompanied by a 95% CI and a corresponding p-value.

#### 3.4.9 SECONDARY OUTCOME 9: Patient-reported insertion pain for initial PIVCs (0-10 numerical rating scale)

Rating scales used to assess reported pain during the insertion of initial PIVCs will be treated as a continuous variable and the mean and SD of the ratings will be calculated per group. The between-group difference will be computed using linear regression, with study group as the main effect. The result will be presented as mean difference along with a 95% CI and a corresponding p-value. If assumptions for the linear model are not met, a quantile regression analysis will be used with the study group as the main effect. The results will be presented as the median difference, along with a 95% CI and a p-value.

#### 3.4.10 SECONDARY OUTCOME 10: Patient-reported satisfaction of PIVC insertion and removal for the initial study PIVCs (0-10 numerical rating scale)

The rating scales for satisfaction regarding the insertion and removal of the first study PIVCs, will be treated as continuous variables. For each study group, the mean and SD of the ratings will be calculated separately.

To assess the between-group difference in satisfaction ratings, linear regression analysis will be conducted, with the study group as the main effect. The outcome of this analysis will be presented as the mean difference, along with a 95% CI and a corresponding p-value.

However, if the assumptions for the linear regression model are not met, a quantile regression analysis will be performed as an alternative. The quantile regression analysis will be used with the study group as the main effect. The result of this analysis will be presented as the median difference, along with a 95% CI and a p-value.

#### 3.4.11 SECONDARY OUTCOME 11: Staff overall satisfaction with the initial PIVC device (0-10 numerical rating scale)

The rating scales for staff overall satisfaction with the initial PIVCs, will be treated as continuous variables. For each study group, the mean and SD of the ratings will be calculated separately. To assess the between-group difference in satisfaction ratings, linear regression analysis will be conducted, with the study group as the main effect. The outcome of this analysis will be presented as the mean difference, along with a 95% CI and a corresponding p-value.

If the assumptions for the linear regression model are not met, a quantile regression analysis will be used as an alternative. The quantile regression analysis will be used with the study group as the main effect. The result of this analysis will be presented as the median difference, along with a 95% CI and a p-value.

#### 3.4.12 SECONDARY OUTCOME 12: Cost analysis

In a subset of seven patients per study arm, the cost of consumables utilized for PIVC insertion will be collected. This data will include the costs associated with the necessary materials and supplies used during the PIVC insertion procedure.

Procedural timings will be observed for ten PIVC insertions (Long, 4; and Short, 3 with ultrasound (US) guidance; Short, 3 without US guidance). The time taken for these insertions will be recorded and subsequently costed based on the "registered nurse" pay scale as per Queensland Health guidelines. This cost calculation will help estimate the labour cost associated with the PIVC insertion procedure.

Furthermore, if any PIVCs require replacement due to device failure, the costs of reinsertion will also be included in the analysis. This accounts for the expenses associated with removing the failed PIVC and inserting a new one.

Additionally, the costs related to any PIVC-related complications, such as bloodstream infections, will be calculated within the specified subset of patients. This allows for an assessment of the financial impact of complications associated with PIVCs.

By considering the costs of consumables, procedural timings, reinsertion costs, and complications, the study aims to comprehensively evaluate the financial aspects associated with PIVC insertions and their potential complications in the specified subset of patients.

## 4. Differences between protocol and statistical analysis plan

The definitions and analyses of the primary and secondary outcomes detailed in this SAP do not differ from the study protocol.

## 5. Trial status

The LEADER trial opened to recruitment in late 2021, however this has been impacted by COVID-19 surges within the health service. The initial SAP was drafted 19/7/2023. Data collection ceased 24/02/2023, and the final database clean and lock occurred on 08/10/2023. This final SAP was finalised after data collection was complete, but data cleaning was ongoing. The analysis of primary and secondary outcomes will be conducted at the completion of the trial and following the approval of this SAP.

## Data Availability

All data produced in the present study are available upon reasonable request to the authors.

## List of abbreviation

BSI: blood stream infection
CI: confidence interval
CVAD: central venous access devices
ID: identification number
IQR: interquartile range
IV: intravenous
PIVC: peripheral intravenous catheter
RCT: randomised controlled trial research nurse
ReN REDCap RR: research electronic data capture risk ratio
SD SAP US: standard deviation statistical analysis plan ultrasound

## Appendix 1. Proposed figures and tables

**Figure 1.**
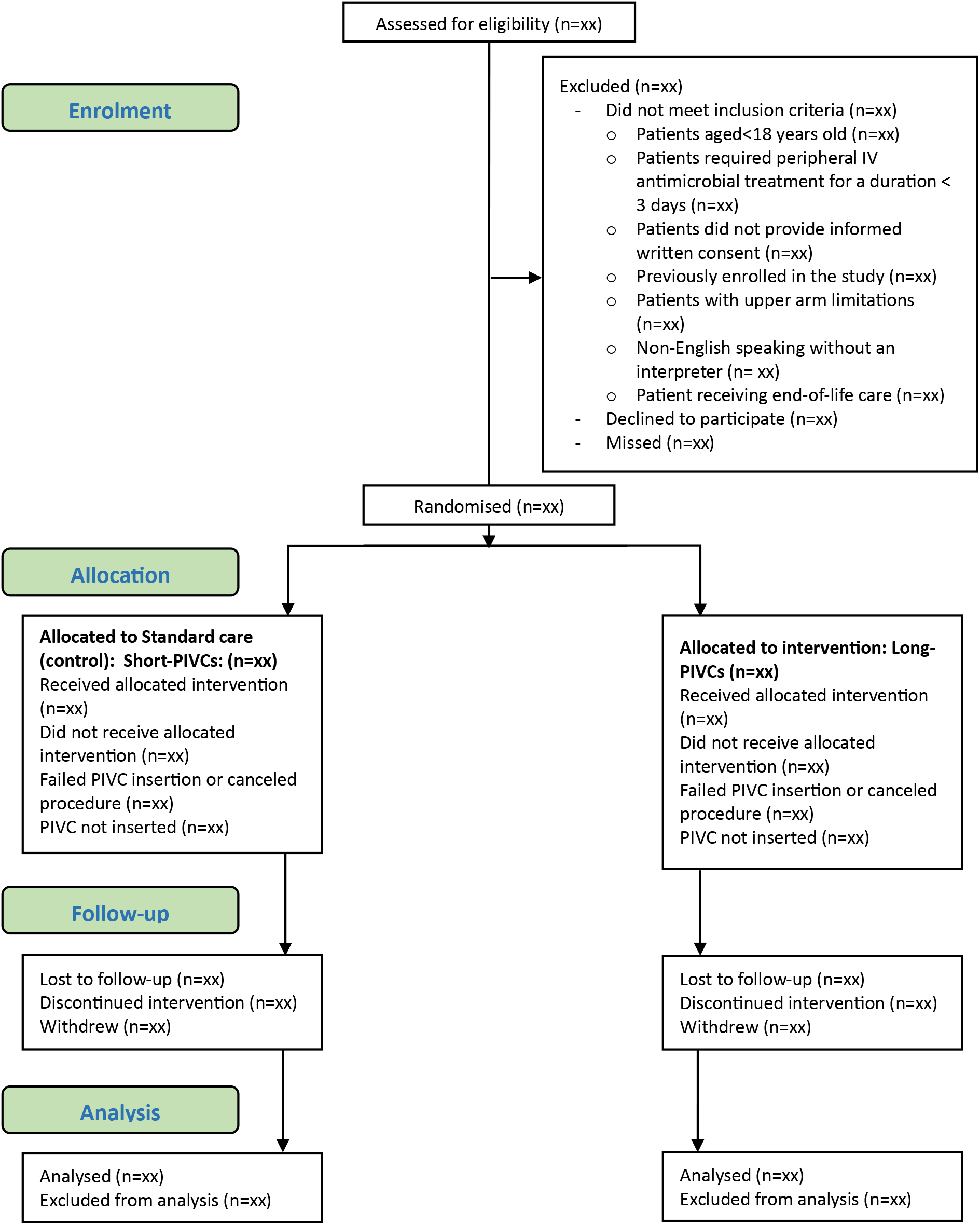
Consort participant flow diagram.

**Table 1.** Data collected by study time point.

| Characteristic | Control condition | Intervention condition |
| --- | --- | --- |
|  | (Short-PIVCs) | (Long-PIVCs) |
|  | Frequency (%) or Mean (SD) or<br>Median (IQR) | Frequency (%) or Mean (SD) or<br>Median (IQR) |
| As per section 3.2.1 | xx (xx) | xx (xx) |

**Table 2.** Device characteristics.

| Characteristic | Control condition | Intervention condition |
| --- | --- | --- |
|  | (Short-PIVCs) | (Long-PIVCs) |
|  | Frequency (%) or Mean (SD) or<br>Median (IQR) | Frequency (%) or Mean (SD) or<br>Median (IQR) |
| As per section 3.2.2 | xx (xx) | xx (xx) |

**Table 3.**
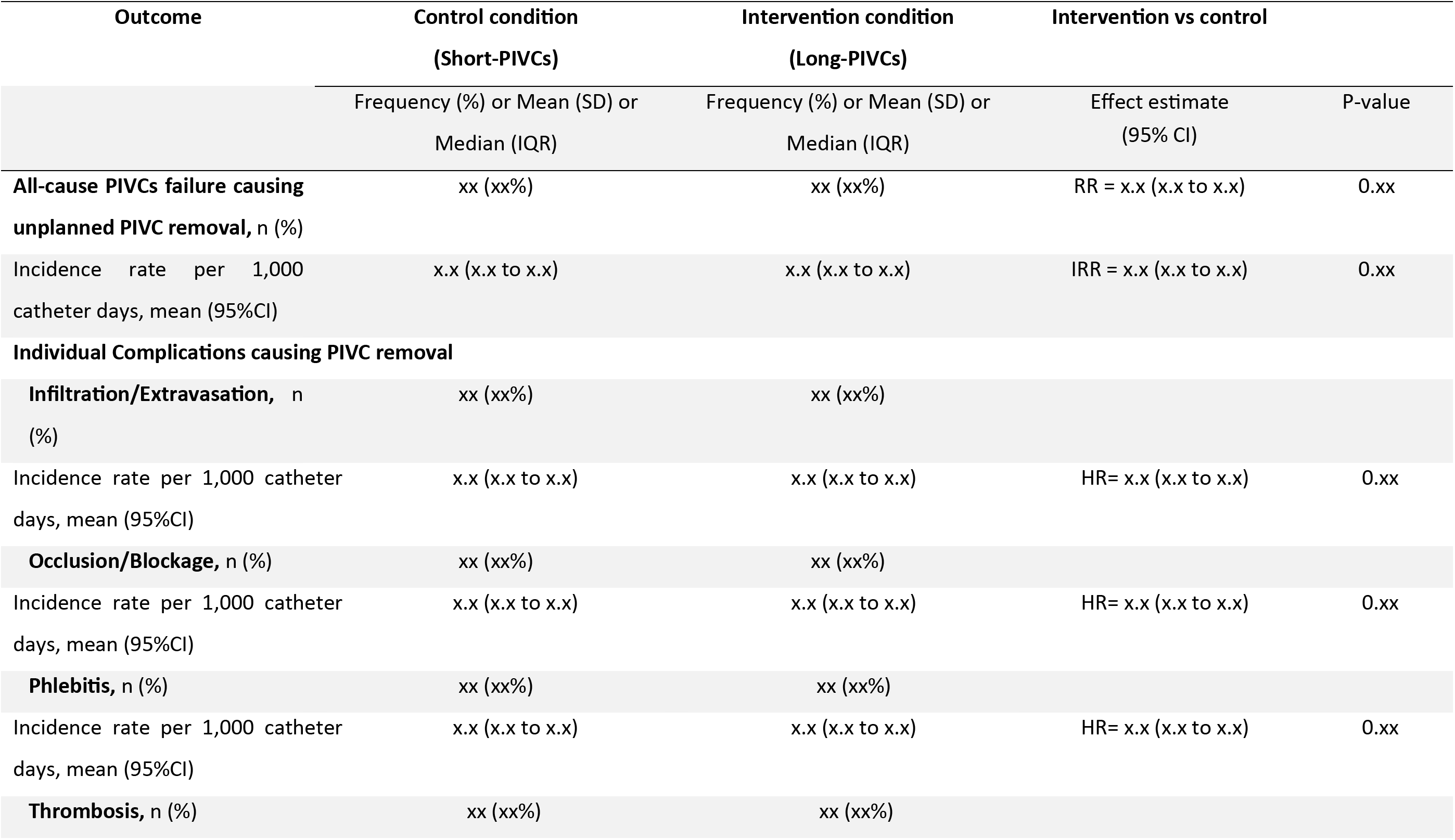
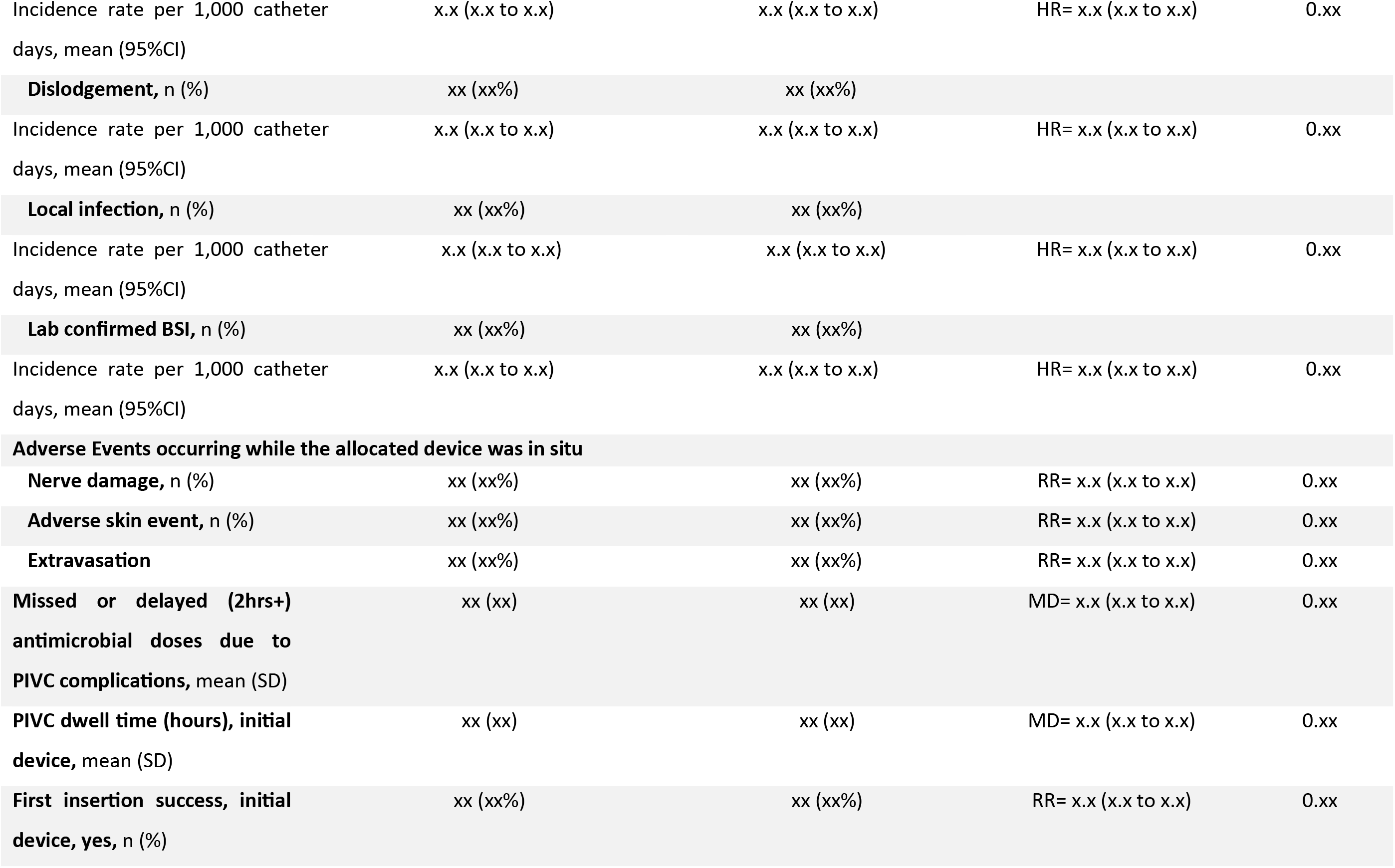
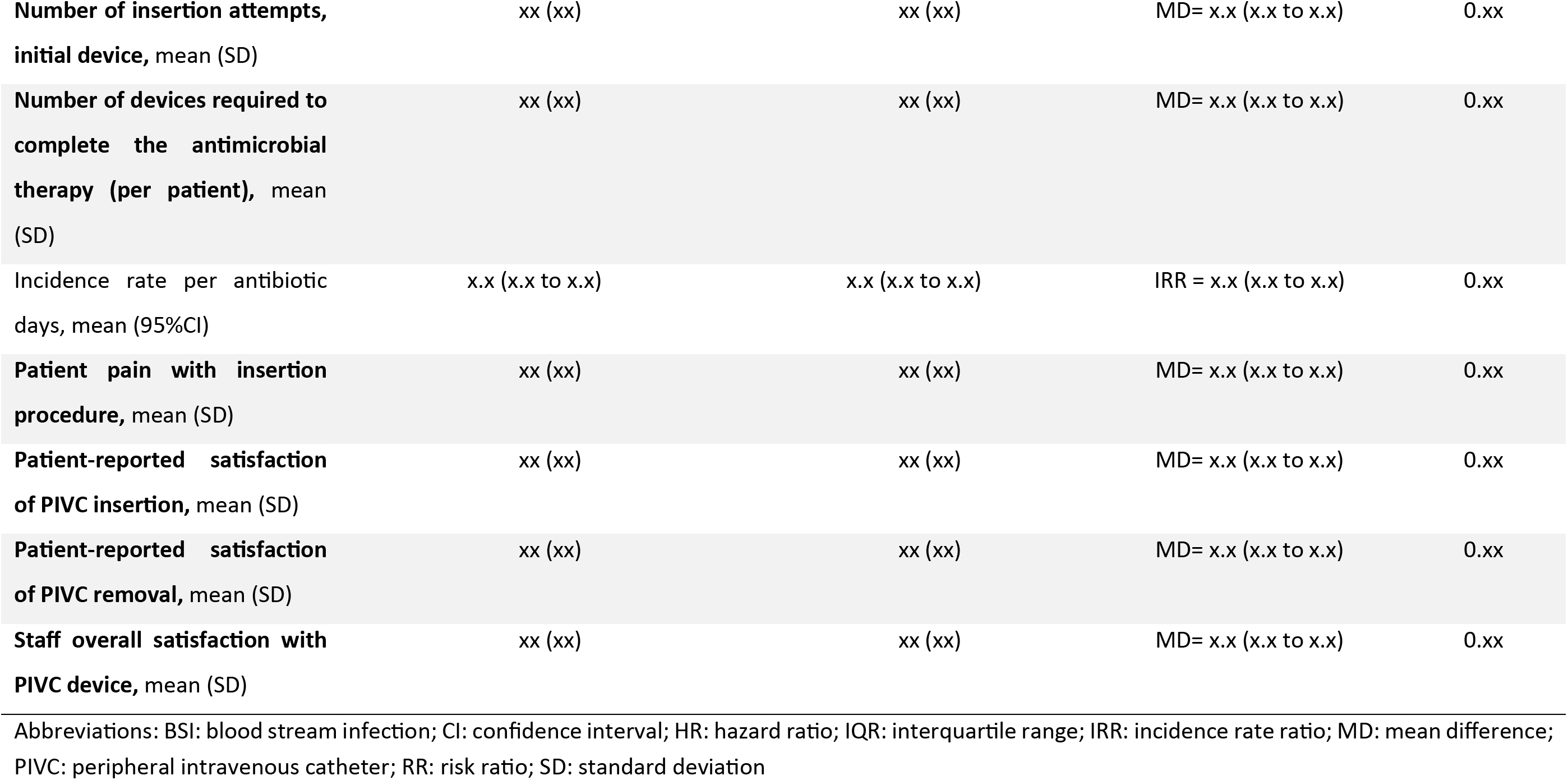
Association between study interventions and study outcome.

**Table 4:**
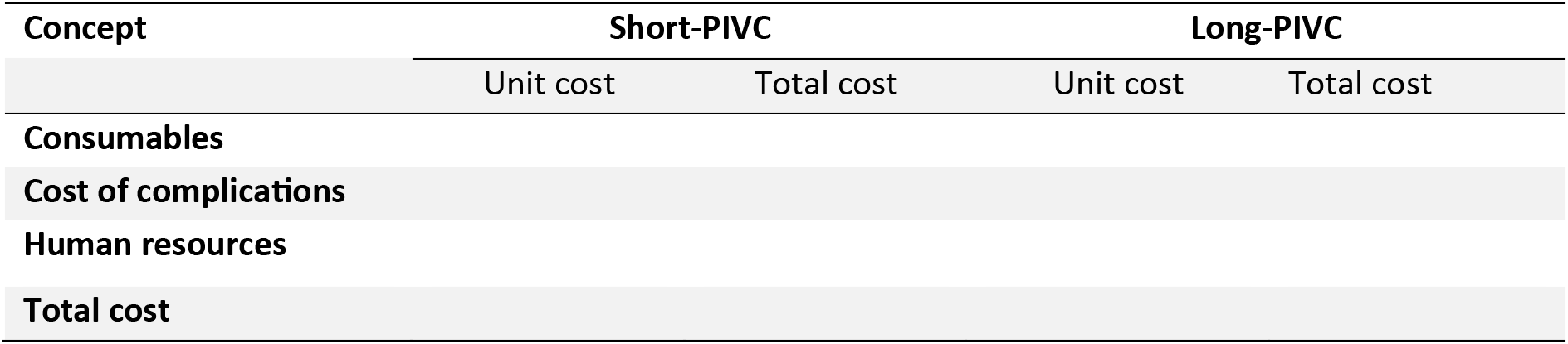
Type and dwell time of subsequent devices used to complete the antimicrobial therapy.

**Table 5:**
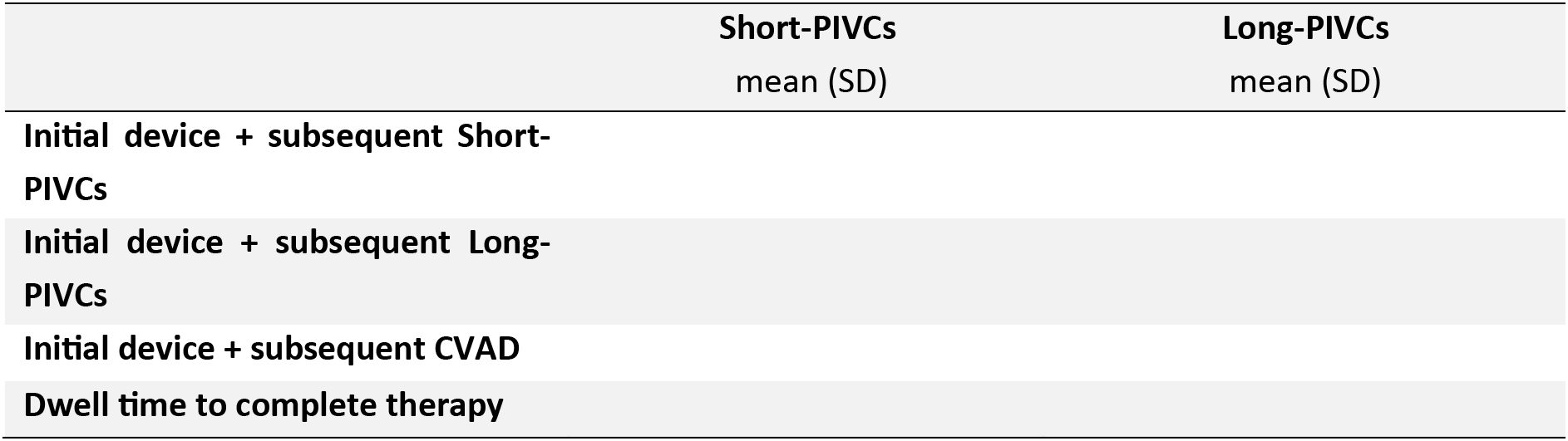
Estimated cost of PIVC use including PIVC-associated complications.

